# Exploring the Intersectionality of Marital Status and Sex on Survival Outcomes in Non-Hodgkin’s Lymphoma

**DOI:** 10.1101/2025.02.27.25322996

**Authors:** Oluwasegun Akinyemi, Mojisola Fasokun, Fadeke Ogunyankin, Akachukwu Eze, Faith Abodunrin, Chisom Ogbuehi, Kakra Hughes, Miriam Michael, Olufunmilayo Abobarin-Aofolaju

## Abstract

**Introduction:** Non-Hodgkin’s Lymphoma (NHL) remains a significant public health concern with notable disparities in survival outcomes. Sex and marital status are two critical social determinants that influence both cancer-specific survival (CSS) and overall survival (OS). Prior studies suggest that women with NHL face poorer survival outcomes compared to men, and marital status modulates survival, with married individuals demonstrating better outcomes. However, the interplay between sex and marital status on NHL survival has not been thoroughly examined.

**Objective:** To investigate the combined effects of sex and marital status on CSS and OS in NHL patients.

**Methodology:** A retrospective cohort study was conducted using SEER registry data spanning 2000 to 2020. Individuals aged 18–85 diagnosed with NHL were included. Cox regression models assessed the interaction between sex and marital status on CSS and OS, adjusting for covariates such as age, race, household income, cancer stage, and treatment modalities. Predicted probabilities of mortality and the differences in sex effects across marital status categories were estimated using interaction terms and linear combinations.

**Results:** A total of 291,608 patients were included, with females accounting for 54.6% of the cohort. Sex disparities were observed across marital status groups. Married women exhibited a significantly higher probability of overall mortality than married men (9.26, 95% CI: 7.61–10.92, p < 0.001). The disparity was most pronounced among single individuals, where women had a 22.93-unit higher probability of overall mortality compared to men (95% CI: 18.49–27.37, p < 0.001). Similarly, widowed women had higher probabilities of mortality than widowed men (13.10, 95% CI: 10.08–16.11, p < 0.001). Cancer-specific mortality followed a similar pattern, with single women experiencing the greatest disparity compared to single men (8.44, 95% CI: 6.21–10.68, p < 0.001). Comparisons of effects between marital status groups revealed that single individuals exhibited a 13.67-unit greater disparity in overall mortality than married individuals (95% CI: 10.26–17.07, p < 0.001).

**Conclusion:** This study underscores the significant influence of sex and marital status on survival outcomes in NHL patients. Women face higher cancer-specific and overall mortality risks across all marital status categories. Marital status modifies these disparities, with marriage conferring a survival advantage, particularly for men. These findings highlight the need for targeted interventions to address sex and social disparities in NHL outcomes, emphasizing the importance of social support and tailored survivorship care.

**Key Points:**

- Women with NHL have higher overall and cancer-specific mortality than men, with the greatest disparity seen in single individuals.
- Marriage reduces survival disparities, benefiting men most, while single and widowed women face the highest mortality risks.

## Introduction

Non-Hodgkin’s Lymphoma (NHL) is a group of hematologic malignancies with a significant public health burden, representing approximately 4% of all cancer diagnoses in the United States.[1–3] The American Cancer Society estimates that in 2024, 80,620 new cases of non-Hodgkin lymphoma (NHL) will be diagnosed in the United States, with approximately 20,140 deaths attributed to the disease.[4] NHL disproportionately affects men, who exhibit higher incidence rates compared to women; however, women with NHL often face worse survival outcomes.[5,6] These disparities in survival have been attributed to a complex interplay of biological, social, and healthcare access factors.[7,8] Beyond the personal toll on patients and families, the economic burden of NHL is immense.[9,10] The total annual cost associated with NHL in the United States, including direct medical expenses and indirect costs such as loss of productivity, exceeds $20 billion.[11] This underscores the critical need for focused efforts to understand and address disparities in NHL outcomes.

The etiology of NHL is multifactorial, with a range of established risk factors contributing to its development. Immunosuppression plays a central role, with conditions such as HIV/AIDS, post-transplant immunosuppression, and autoimmune diseases significantly increasing the risk of NHL. Infections such as Epstein-Barr virus (EBV) and Helicobacter pylori have also been implicated in certain subtypes of the disease.[12,13] Additionally, environmental exposures to chemicals, such as pesticides and herbicides, have been associated with NHL risk.[14,15] Demographic factors, including advanced age, male sex, and Caucasian race, further compound the risk.[12,16] While these factors have been well-studied, understanding their interaction with psychosocial determinants, such as marital status, remains limited.

The role of sex and marital status in determining survival outcomes has been extensively studied in various cancers and other clinical conditions.[17,18] In breast and colorectal cancers, women have been shown to have better survival rates than men, potentially due to differences in tumor biology and healthcare-seeking behaviors.[5,19,20] However, in hematologic malignancies like NHL, women often exhibit worse outcomes[21], which may be influenced by differences in treatment access, adherence, and immune response. Similarly, marital status has emerged as a significant determinant of survival across numerous cancers.[17] Married individuals consistently demonstrate better survival compared to their unmarried counterparts.[17] This phenomenon is attributed to the emotional, logistical, and financial support provided by a spouse, which facilitates early diagnosis, timely treatment, and adherence to care plans.[18,22,23]

The protective effect of marriage has been observed across both genders, though it appears more pronounced in men.[17] Married men often experience substantial survival benefits in cancers such as lung, prostate, and gastrointestinal malignancies compared to single, divorced, or widowed men.[17] In contrast, while married women also experience improved survival compared to unmarried women, the magnitude of this benefit is often less than that seen in men.[18,24] This discrepancy suggests that marriage may confer different types or levels of support for men and women, with women potentially serving as caregivers even within marital relationships, which may limit their ability to fully benefit from spousal support.

Despite these findings, the intersectionality of sex and marital status on survival outcomes in NHL remains underexplored. While some studies have highlighted the role of marital status[24] and sex[5] independently, few have examined how these two factors interact to influence survival outcomes. Moreover, little is known about how these relationships vary across specific marital status categories, such as divorced or widowed individuals, and whether sex disparities persist within these groups. To address this gap, our study aims to investigate the combined impact of sex and marital status on both cancer-specific and overall mortality in NHL. By leveraging the SEER registry’s comprehensive data, we aim to provide a nuanced understanding of these interactions, ultimately informing targeted interventions to reduce disparities and improve survival outcomes for NHL patients.

## Methodology

### Study Design and Population

This retrospective cohort study analyzed data from the Surveillance, Epidemiology, and End Results (SEER) cancer registry to evaluate the impact of sex and marital status on cancer-specific and overall mortality among patients diagnosed with Non-Hodgkin’s Lymphoma (NHL). The SEER registry[25] is a nationally representative database that collects comprehensive cancer incidence and survival data from population-based cancer registries covering approximately 48% of the U.S. population.[26]

The study population included individuals aged 18 years and older who were diagnosed with NHL between January 1, 2000, and December 31, 2020. Patients with missing data on sex, marital status, cancer stage, or treatment modalities were excluded from the analysis, as were those with incomplete follow-up or mortality information.

### Data Source

The SEER cancer registry provided detailed information on demographic characteristics (e.g., age, sex, race/ethnicity, and marital status), tumor-specific factors (e.g., histology, stage at diagnosis), treatment modalities (e.g., surgery, chemotherapy, radiotherapy), and survival outcomes.[25] Mortality data were derived from linked national death records, allowing precise determination of NHL-specific mortality and overall mortality.[27]

### Study Variables

The study focused on two primary outcomes: cancer-specific mortality and overall mortality. Cancer-specific mortality was defined as deaths attributed directly to NHL, as recorded in the SEER mortality data. This measure allowed for precise evaluation of mortality directly associated with the disease. Overall mortality, on the other hand, was defined as deaths from any cause occurring during the study period. This broader measure provided insights into the general survival of the population, accounting for both cancer-related and unrelated deaths.

### Variable of Interest

The key explanatory variables in this study included sex and marital status, as well as their interaction. Sex was coded as a binary variable, with males serving as the reference group (male = 0, female = 1). Marital status was categorized into five distinct groups: married, single (never married), widowed, divorced, and separated. These categories enabled a nuanced analysis of how marital status influences survival outcomes. To assess the combined effects of sex and marital status on mortality, an interaction term between these variables was included in the analysis. This interaction term was critical for examining whether the sex disparities in mortality varied across different marital status categories.

### Covariates

Several covariates were incorporated to control for potential confounders and ensure robust estimates. Age at diagnosis was treated as a continuous variable to account for its direct influence on survival outcomes. Cancer stage was categorized based on the Ann Arbor staging system, ranging from Stage I (localized disease) to Stage IV (advanced disease with systemic involvement). Race and ethnicity were categorized into five groups: non-Hispanic White, non-Hispanic Black, Hispanic, Asian/Pacific Islander, and others. These categories allowed for the exploration of racial and ethnic disparities in mortality and survival outcomes.

Treatment modalities were included as binary indicators, reflecting whether patients received surgery, chemotherapy, and/or radiotherapy during their treatment course. These variables captured the influence of specific therapeutic interventions on survival outcomes. Socioeconomic status (SES) was proxied using area-level household median income data, categorized into quartiles. This measure provided insight into how economic factors influenced survival outcomes, recognizing that access to care and treatment adherence often vary by SES.

By including these variables, the study aimed to comprehensively assess the intersectionality of sex, marital status, and other demographic and clinical factors on NHL-specific and overall survival outcomes.

### Statistical Analysis

Descriptive statistics were used to summarize the cohort’s demographic and clinical characteristics. Categorical variables were reported as frequencies and percentages, while continuous variables were summarized using means and standard deviations or medians and interquartile ranges, depending on the data distribution. Group differences were assessed using chi-square tests for categorical variables and t-tests for continuous variables.

### Survival Analysis

Cox proportional hazards regression models were employed to estimate hazard ratios (HRs) for cancer-specific and overall mortality. The models included sex, marital status, and their interaction as key predictors, adjusting for age, stage at diagnosis, race/ethnicity, treatment modalities, and socioeconomic status. The proportional hazards assumption was tested using Schoenfeld residuals.

### Marginal Effects and Interaction Testing

To further evaluate the sex and marital status interaction, predicted probabilities of cancer-specific and overall mortality were computed using the margins command in Stata (VERSION 16). Linear combinations of coefficients (lincom) were used to:

1. Compare sex effects within each marital status category (e.g., married women vs. married men).
2. Examine differences in sex effects between marital status categories (e.g., single vs. married).

### Sensitivity Analyses

Sensitivity analyses were conducted to ensure the robustness of the findings. These included:

1. Restricting the cohort to patients aged <65 years to evaluate the consistency of results in younger populations.
2. Employing stratified Cox models for scenarios where the proportional hazards assumption was not met.

All analyses were performed using Stata (version 16.0). Statistical significance was determined at a two-sided p-value < 0.05.

## Ethical Considerations

The SEER registry provided de-identified data; therefore, informed consent or IRB approval was not required. The study adhered to the principles outlined in the Declaration of Helsinki and followed all applicable ethical guidelines and regulations for research involving human data.

## Results

Table 1 presents the baseline characteristics of 291,608 patients diagnosed with Non-Hodgkin’s Lymphoma (NHL) between 2000 and 2020, stratified by sex. The mean age of the cohort was 64.9 years (±14.9), with males being significantly older than females (66.0 ± 14.7 years vs. 63.9 ± 15.0 years, p < 0.001). The age distribution showed that a larger proportion of population were greater than 65 years of age. Females were more prevalent in the < 45 years group (11.3% vs. 9.0%, p < 0.001), whereas males were more likely to be 65 years or older (59.0% vs. 53.5%, p < 0.001). Regarding race and ethnicity, the majority of the population was White (73.4%), with similar proportions across sex.

**Table 1:** Baseline Characteristics of Non-Hodgkin’s Lymphoma Patients Stratified by Gender ( SEER Registry 2000-2020) Table 1 presents the baseline demographic, clinical, and treatment characteristics of 213,250 patients diagnosed with Non-Hodgkin’s Lymphoma (NHL) between 2000 and 2020, stratified by gender (male vs. female).

| Variable | Total Population | Male | Female | Statistics | p-value |
| --- | --- | --- | --- | --- | --- |
|  | (N=291,608) | (N=132,435 (45.4%)) | (N=159,173 (54.6%)) |  |  |
| Age, year, Mean ± SD | 64.9 + 14.9 | 66.0 + 14.7 | 63.9 + 15.0 | t = 36.14 | < 0.001 |
| Age (yrs.) | | | | $\chi^2$ =958.554 | <0.001 |
| <45yRS | 29,131 (10.3%) | 11,494 (9.0%) | 17,637 (11.3%) |  |  |
| 45-64Yrs. | 95,735 (33.8%) | 40,898 (32.0%) | 54,837 (35.2%) |  |  |
| ≥65Yrs. | 158,704 (56.0%) | 75,362 (59.0%) | 83,342 (53.5%) | | |
| Race/Ethnicity | | | | $\chi^2$= 44.1862 | <0.001 |
| White | 213,250 (73.4%) | 96,140 (72.9%) | 117,110 (73.9%) |  |  |
| Black | 20,754 (7.1%) | 9,466 (7.2%) | 11,288 (7.1%) |  |  |
| Hispanic | 35,498 (12.2%) | 16,569 (12.6%) | 18,929 (11.9%) |  |  |
| Other | 21,034 (7.2%) | 9,799(7.4%) | 11,235 (7.1%) |  |  |
| Grade | | | | $\chi^2$= 148.6729 | <0.001 |
| Stage I | 68,075(47.6 %) | 32,588 (48.9%) | 35,487 (46.5%) |  |  |
| Stage II | 41,443 (29.0%) | 19,358 (29.1%) | 22,085 (28.9%) |  |  |
| Stage III | 11,501 (8.1%) | 5,159 (7.8%) | 6,342 (8.3%) |  |  |
| Stage IV | 21,891 (15.3%) | 9,500 (14.3%) | 12,391 (16.2%) |  |  |
| Marital Status | | | | $\chi^2$=181.04 | <0.001 |
| Single | 46,066 (15.8%) | 18,869 (14.3%) | 27,197(17.1%) |  |  |
| Married | 174,481 (59.3%) | 65,752 (49.7%) | 108,729 (68.3%) |  |  |
| Widowed | 44,192 (15.2%) | 33,304 (25.2%) | 10,888 (6.8%) |  |  |
| Divorced | 24,258<br>(8.3%) | 13,294 (10.0%) | 10,964 (6.9%) |  |  |
| Separated | 2,611<br>(0.9%) | 1,216 (0.9%) | 1,395 (0.9%) |  |  |
| Metropolitan Status | | | | $\chi^2=$<br>0.0001 | 0.992 |
| Metropolitan | 256,606<br>(88.1%) | 116,536<br>(88.1%) | 140,070<br>(88.1%) |  |  |
| non-Metropolitan | 34,815<br>(12.0%) | 15,812 (12.0%) | 19,003 (12.0%) |  |  |
| Household Median<br>Income | | | | $\chi^2=22.66$<br>30 | <<br>0.001 |
| Income I | 25,542<br>(8.8%) | 11,681 (8.8%) | 13,861 (8.7%) |  |  |
| Income II | 38073<br>(13.1%) | 17,552 (13.3%) | 20,521 (12.9%) |  |  |
| Income III | 99,693<br>(34.2%) | 45540 (34.4%) | 54,153 (34.0%) |  |  |
| Income IV | 128,276<br>(44.0%) | 57,650 (43.5%) | 70,626 (44.4%) |  |  |
| Treatment<br>Modalities |  |  |  |  |  |
| Surgery, n (%) | | | | $\chi^2=$<br>31.7039 | 0.023 |
| None/Unknown | 204,026<br>(72.7%) | 91,962 (72.2%) | 112,064<br>(73.2%) |  |  |
| Yes | 76,525<br>(27.3%) | 35,402 (27.8%) | 41,123 (26.8%) |  |  |
| Chemotherapy, n<br>(%) | | | | $\chi^2=$<br>704.7761 | <0.00<br>1 |
| None/Unknown | 125,086<br>(42.9%) | 60,341 (45.6%) | 5,367 (38.5%) |  |  |
| Yes | 166,522<br>(57.1%) | 72,094 (54.4%) | 8582(61.5%) | $\chi^2=$ 39.18 | <0.00<br>1 |
| Radiotherapy , n(%) | | | | $\chi^2=$<br>10.7010 | |
| None/Unknown | 241,972<br>(83.8%) | 109,488<br>(83.5%) | 132,484<br>(84.0%) |  |  |
| Radiotherapy | 46,813<br>(16.2%) | 21,567<br>(16.5%) | 25,246 (16.0%) | $\chi^2=$<br>69.124 | <0.00<br>1 |
| 5-Year Survival<br>(CSS) | 27255<br>(72.1%) | 11,479 (74.0%) | 15,776 (70.6%) |  |  |
| 5-Year Survival<br>(OS) | 47096<br>(60.7%) | 19,991 (63.3%) | 27,105 (58.6%) |  |  |

Clinical characteristics showed significant differences by sex. A higher proportion of males were diagnosed with early-stage disease (Stage I: 48.9% vs. 46.5%, p < 0.001), whereas females were more likely to present with advanced disease (Stage IV: 16.2% vs. 14.3%, p < 0.001). Marital status also varied significantly; married individuals constituted the largest group overall (59.3%), but the proportion was markedly higher among females (68.3%) than males (49.7%, p < 0.001). Widowed status was more common among males (25.2% vs. 6.8%), reflecting sex differences in life expectancy and social support dynamics.

Treatment patterns and survival outcomes revealed notable disparities. Females were more likely to receive chemotherapy (61.5% vs. 54.4%, p < 0.001), while the use of surgery and radiotherapy was comparable between the sexes. Despite receiving more chemotherapy, females had lower 5-year cancer-specific survival (CSS) rates (70.6% vs. 74.0%, p < 0.001) and overall survival (OS) rates (58.6% vs. 63.3%, p < 0.001) compared to males. These differences suggest that underlying biological, social, and healthcare-related factors may contribute to the observed sex disparities in survival outcomes among NHL patients.

Table 2 highlights the predicted probabilities of overall mortality by sex (female vs. male) across different marital status categories. The results demonstrate significant sex disparities in mortality probabilities within each marital status group (p < 0.001 for all comparisons).

**Table 2:** Predicted Probabilities of Overall mortality by Gender (Female vs. Male) and Marital Status. Table 2 presents the predicted probabilities of overall mortality stratified by gender (female vs. male) and marital status. The table displays the coefficients (Coef.), standard errors (Std. Err.), z-values (z), p-values (P>z), and 95% confidence intervals (95% CI) for the gender effect within each marital status category. Statistically significant differences (p < 0.001) indicate variations in the gender effect on overall mortality across marital status groups.

|  | **Coef.** | **Std. Err.** | **z** | **P>z** | **95% CI** |  |
| --- | --- | --- | --- | --- | --- | --- |
| Married | 9.264 | 0.847 | 10.940 | 0.000 | 7.605 | 10.924 |
| Single | 22.930 | 2.266 | 10.120 | 0.000 | 18.490 | 27.371 |
| Widowed | 13.097 | 1.539 | 8.510 | 0.000 | 10.081 | 16.112 |
| Divorced | 17.124 | 1.976 | 8.670 | 0.000 | 13.251 | 20.997 |
| Separated | 19.070 | 5.325 | 3.580 | 0.000 | 8.633 | 29.507 |

Specifically, married women had a 9.26-unit higher predicted probability of mortality compared to married men (95% CI: 7.61–10.92, p < 0.001). The disparity was more pronounced among single individuals, where women had a 22.93-unit higher predicted probability of mortality compared to single men (95% CI: 18.49–27.37, p < 0.001). Similarly, widowed women exhibited a 13.10-unit higher mortality probability than widowed men (95% CI: 10.08–16.11, p < 0.001), while divorced women showed a 17.12-unit higher probability compared to divorced men (95% CI: 13.25–21.00, p < 0.001). Among separated individuals, women had a 19.07-unit higher predicted probability of mortality compared to men (95% CI: 8.63–29.51, p < 0.001).

Table 3 presents the comparison of sex effects on overall mortality between single and married individuals. The coefficient of 13.67 (95% CI: 10.26–17.07, p < 0.001) represents the difference in the sex effect on overall mortality between the two marital status categories. This result indicates that the disparity in overall mortality between females and males is significantly greater among single individuals compared to married individuals.

**Table 3:** Comparison of Gender Effects on Overall mortality Across Marital Status Categories (Single vs. Married) Table 3 displays the comparison of gender effects on overall mortality between single and married individuals. The table includes the coefficient (Coef.), standard error (Std. Err.), z-value (z), p-value (P>z), and 95% confidence interval (95% CI). The coefficient represents the difference in the gender effect on overall mortality between single and married individuals, with a statistically significant result (p < 0.001) indicating that the gender disparity is greater among single individuals compared to married individuals.

|  | **Coef.** | **Std. Err.** | **z** | **P>z** | **95% CI** |  |
| --- | --- | --- | --- | --- | --- | --- |
| Single vs. Married. | 13.666 | 1.737 | 7.870 | 0.000 | 10.262 | 17.070 |

The statistically significant finding suggests that single women experience a disproportionately higher mortality risk relative to single men when compared to the sex disparity observed among married individuals. This highlights the potential protective effect of marriage in mitigating sex-based disparities in overall mortality.

Table 4 presents the predicted probabilities of cancer-specific mortality by sex (female vs. male) across marital status categories. The findings reveal statistically significant sex disparities in cancer-specific mortality probabilities within all marital status groups (p < 0.001 for all comparisons).

**Table 4:** Predicted Probabilities of Cancer-specific mortality by Gender (Female vs. Male) and Marital Status. Table 4 presents the predicted probabilities of cancer-specific mortality stratified by gender (female vs. male) and marital status. The table displays the coefficients (Coef.), standard errors (Std. Err.), z-values (z), p-values (P>z), and 95% confidence intervals (95% CI) for the gender effect within each marital status category. Statistically significant differences (p < 0.001) indicate variations in the gender effect on cancer-specific mortality across marital status groups.

|  | **Coef.** | **Std. Err.** | **z** | **P>z** | **95% CI** |  |
| --- | --- | --- | --- | --- | --- | --- |
| Married | 3.024 | 0.395 | 7.660 | 0.000 | 2.250 | 3.798 |
| Single | 8.444 | 1.142 | 7.390 | 0.000 | 6.205 | 10.683 |
| Widowed | 4.023 | 0.741 | 5.430 | 0.000 | 2.571 | 5.474 |
| Divorced | 5.085 | 0.887 | 5.730 | 0.000 | 3.347 | 6.823 |
| Separated | 6.676 | 2.553 | 2.620 | 0.000 | 1.673 | 11.680 |

Married women had a 3.02-unit higher predicted probability of cancer-specific mortality compared to married men (95% CI: 2.25–3.80, p < 0.001). Among single individuals, the disparity was more pronounced, with women exhibiting an 8.44-unit higher probability of cancer-specific mortality compared to men (95% CI: 6.21–10.68, p < 0.001). Widowed women had a 4.02-unit higher predicted probability of cancer-specific mortality than widowed men (95% CI: 2.57–5.47, p < 0.001), while divorced women showed a 5.09-unit higher probability compared to divorced men (95% CI: 3.35–6.82, p < 0.001). Among separated individuals, women had the largest disparity, with a 6.68-unit higher predicted probability of cancer-specific mortality compared to men (95% CI: 1.67–11.68, p < 0.001).

Table 5 presents the comparison of sex effects on cancer-specific mortality between single and married individuals. The coefficient of 5.42 (95% CI: 3.65–7.19, p < 0.001) represents the difference in the sex effect on cancer-specific mortality between the two marital status categories. This result indicates that the disparity in cancer-specific mortality between females and males is significantly greater among single individuals compared to married individuals.

**Table 5:** Comparison of Gender Effects on cancer-specific mortality Across Marital Status Categories (Single vs. Married) Table 5 displays the comparison of gender effects on cancer-specific mortality between single and married individuals. The table includes the coefficient (Coef.), standard error (Std. Err.), z-value (z), p-value (P>z), and 95% confidence interval (95% CI). The coefficient represents the difference in the gender effect on cancer-specific mortality between single and married individuals, with a statistically significant result (p < 0.001) indicating that the gender disparity is greater among single individuals compared to married individuals

|  | Coef. | Std. Err. | z | P>z | 95% CI |  |
| --- | --- | --- | --- | --- | --- | --- |
| **Single vs. Married.** | 5.420 | 0.902 | 6.010 | 0.000 | 3.652 | 7.188 |

## Discussion

This study reveals significant disparities in survival outcomes based on sex and marital status. The findings show that women, irrespective of their marital status, consistently exhibited higher cancer-specific and overall mortality rates following an NHL diagnosis. This disparity translates to lower NHL-specific and overall survival rates for women compared to men. The results further highlight that, across all marital status groups, women had persistently higher mortality rates than men. These observations suggest a complex interplay of biological, social, and systemic factors influencing survival outcomes in NHL.

One of the most striking findings is the consistent survival disadvantage faced by women across all marital status categories. Biological differences, such as variations in immune response, tumor biology, or hormonal influences, may partly explain these disparities.[6] Additionally, systemic barriers, including unequal access to care, delayed diagnosis, and challenges in treatment adherence, may disproportionately affect women.[28] Beyond biological and healthcare system factors, social determinants of health likely play a critical role.[29,30] Women often face greater caregiving responsibilities, economic challenges, and social pressures that could negatively impact their ability to prioritize their own health and adhere to treatment protocols.[31,32] These findings underscore the need for a comprehensive approach to address the multifaceted challenges faced by women with NHL.

Marriage was found to have a protective effect on survival, particularly among men. Married men exhibited the lowest cancer-specific and overall mortality rates, suggesting that marital status plays a significant role in improving survival outcomes. The benefits of marriage may stem from increased emotional, physical, and logistical support provided by a spouse, which can encourage timely healthcare-seeking behavior and treatment adherence.[33] Marriage may also confer financial stability, which facilitates better access to healthcare resources.[18,22] Interestingly, while married women also experienced improved survival compared to their single, divorced, or widowed counterparts, they continued to have higher mortality rates than married men.[22] This indicates that the protective effect of marriage may not be equally distributed between sexes. Married women may still face unique challenges, such as balancing caregiving roles with their own health needs, which could limit their ability to fully benefit from the support structures of marriage.[18,34]

The study also highlights significant disparities within specific marital status categories. Single women experienced the highest mortality rates, reflecting the compounded challenges faced by women without spousal support. Widowed, divorced, and separated women also showed higher mortality rates compared to their male counterparts, although to a lesser extent than single women. These findings emphasize the critical role of social support in improving survival outcomes.[17] The absence of a partner may exacerbate vulnerabilities, particularly for women, who may have fewer resources or less social support in navigating their illness.[17] In contrast, single men, while also disadvantaged compared to married men, had better survival outcomes than single women, further underscoring the gendered nature of these disparities.[35]

The persistent sex disparities in survival outcomes, even among married individuals, highlight the need for targeted interventions. Women appear to face unique challenges that are not entirely mitigated by the presence of a spouse. Tailored interventions that address these challenges are essential. For example, improving access to care, providing psychosocial support, and addressing systemic barriers such as financial strain and caregiving responsibilities could help reduce these disparities. Additionally, healthcare providers should consider the role of social support systems beyond marriage and encourage community-based interventions that provide support for single, widowed, and divorced individuals, particularly women.

These findings also have implications for research and public health practice. Future research should explore the biological, hormonal, and psychosocial factors that contribute to these sex disparities in NHL outcomes. Clinical trials should prioritize sex-specific analyses to ensure that treatments are equally effective for men and women. Furthermore, public health initiatives should aim to raise awareness of these disparities and advocate for systemic changes that reduce inequities in care delivery and access.

Despite its strengths, including a large sample size and detailed marital status categorization, this study has some limitations. The observational nature of the study may be subject to residual confounding from unmeasured variables such as socioeconomic status or comorbidities. Additionally, marital status was measured at diagnosis, and subsequent changes in marital status, which could influence survival outcomes, were not captured.

In conclusion, this study underscores the significant sex disparities in NHL-specific and overall survival, with women consistently facing higher mortality rates than men across all marital status categories. While marriage was associated with improved survival outcomes overall, the protective effect was more pronounced for men than women. These findings highlight the need for targeted interventions, systemic reforms, and sex-specific research to address the barriers contributing to these disparities. Addressing these challenges is critical to improving survival outcomes for all NHL patients, particularly women.

## Data Availability

All relevant data are within the manuscript and its Supporting Information files.

## Notes

### Competing Interest Statement

The authors have declared no competing interest.

### Funding Statement

The author(s) received no specific funding for this work.

### Author Declarations

The SEER registry provided de-identified data therefore, informed consent or IRB approval was not required.

